# Quantitative Plasma Proteomics Identifies Metallothioneins as a Marker of Acute-on-Chronic Liver Failure Associated Acute Kidney Injury

**DOI:** 10.1101/2022.05.30.22275767

**Authors:** Pragyan Acharya, Rohini Saha, Javed Ahsan Quadri, Saba Sarwar, Maroof Ahmad Khan, Hem Chandra Sati, Nidhi Gauniyal, Ahmadullah Shariff, Shekhar Swaroop, Piyush Pathak, Shalimar

## Abstract

**Objective:** Acute kidney injury (AKI) considerably increases the risk of short-term mortality in acute-on-chronic liver failure (ACLF) but predicting AKI is not possible with existing tools. Our study aimed at *de novo* discovery of AKI biomarkers in ACLF.

**Design:** This observational study had two phases- (A) Discovery phase in which quantitative proteomics was carried-out with day-of-admission plasma from ACLF patients who initially had no-AKI but progressed to AKI (n=10) or did not progress to AKI (n=9) within 10 days of admission and, (B) Validation phase in which selected biomarkers from the discovery phase were validated by ELISA in a larger set of ACLF plasma samples (n=93) followed by sub-group analyses.

**Results:** Plasma proteomics revealed 56 differentially expressed proteins in ACLF patients who progressed to AKI vs those who did not. The metallothionein protein-family was upregulated in patients who progressed to AKI and was validated by ELISA as significantly elevated in both- (i) ACLF AKI vs no-AKI (p-value≤0.0001) and (ii) progression to AKI vs no-progression to AKI (p-value≤0.001). AUROC for AKI vs no-AKI was 0.786 (p-value ≤0.001) and for progression to AKI vs no-progression to AKI was 0.7888 (p-value ≤0.001). Kaplan-Meier analysis revealed that ACLF patients with plasma MT concentration >5.83 ng/mL (cut-off defined at 80% specificity and 80% sensitivity) had a high probability of developing AKI by day 7 (p-value ≤0.0001). High expression of metallothionein genes was found in post-mortem liver biopsies of ACLF patients.

**Conclusion:** Day-of-admission measurements of plasma metallothionein can act as predictive biomarkers of AKI in ACLF.

**Key Messages:** *What is already known about this subject?:* In ACLF, AKI is a key event that significantly increases the risk of mortality. Therefore, the ability to predict AKI in an ACLF patient on the day-of-diagnosis or the day-of-admission would be beneficial in order to tailor management of such patients. The existing gold standards for detection of AKI are serum creatinine and urea which have not been proven to be accurate and consistent in the prediction of AKI. Systematic discovery studies using high throughput approaches aimed at de novo discovery of predictive AKI biomarkers have not been carried out.

*What are the new findings?:* In a prospective discovery study, day-of-admission plasma samples were subjected to quantitative proteomics from ACLF patients who initially did not present with AKI but either progressed to AKI or did not progress to AKI within a follow-up period of 10 days. 56 differentially expressed proteins plasma proteins were found in the ACLF patients who progressed to AKI as compared to those who did not. Metallothionein family of proteins were overrepresented in the ACLF progression to AKI group. ELISA based validation in a larger ACLF cohort revealed a significant elevation of plasma metallothioneins levels in the day-of-admission plasma of ACLF patients who progressed to AKI compared with those who did not. Plasma metallothionein was elevated also in a cross-sectional analysis of ACLF patients who had clinically diagnosed AKI at admission compared with those that did not. This suggested a strong association of plasma metallothionein with both- the presence of AKI and, the progression of ACLF patients to AKI. Biomarker performance statistics revealed that the probability of developing AKI within 7 days for an ACLF patient significantly increases above a cut-off of 5.83 ng/mL plasma metallothionein concentration on the day-of-admission.

*How might it impact on clinical practice in the foreseeable future?:* Plasma metallothionein levels can be easily measured through ELISA and therefore, can be converted to a bedside day-of-admission test for ACLF patients in order to evaluate their risk of developing AKI. This may assist clinicians to tailor their management strategies and closely monitor the renal function of such patients during their management of ACLF thereby improving their chances of recovery.

## INTRODUCTION

Acute-on-Chronic Liver Failure (ACLF) is a complex liver disease with a very high short-term mortality of ∼50%^1,2^. The hallmarks of ACLF are- systemic inflammation, innate immune dysfunction and extra-hepatic multiple organ failure (MOF)^3, 4^. The chief reason for mortality in ACLF is MOF^5^. Among MOF, acute kidney injury (AKI) is the central event that changes the course of the disease leading to poor patient outcomes^6,7^

Recent evidencesuggests that the mechanism of ACLF associated AKI is different from hepatorenal syndrome (HRS) and involves an increase in inflammatory markers such as TLR4 and caspase 3 in kidney biopsies from ACLF ^6,8,9^. Reported observations show that ACLF-AKI is associated with acute tubular necrosis, a greater reversibility along with rapid progression suggesting therefore that prediction and prevention of AKI in ACLF is a top priority to prolong the window of recovery for the patient, during which reversal of AKI can take place and the risk of mortality can reduce^8,9^. However, currently, there are no predictive markers of AKI in ACLF, and neither are therapeutic targets for the prevention of AKI known.

The human plasma is an ideal source of biomarkers due to two reasons- (i) it is the source of the most comprehensive human proteome representative of plasma proteins, tissue leakage markers, as well as markers emanating out of diseased cells and, (ii) the ease of plasma collection and preparation using methods that can be uniformly followed all over the world. Our study aimed to identify plasma proteome based molecular changes in ACLF patients who progress to AKI as compared to ACLF patients who did not progress to AKI. Towards this, the objectives of our study were- (i) To identify plasma proteome changes in day-of-admission samples derived from ACLF patients who did not present with AKI initially but who progressed to AKI within 10 days of admission as compared to those who did not and, (ii) to validate selected plasma protein markers in a larger cohort of ACLF patients and carry out further sub-group analyses in them.

We found that quantitative changes in plasma proteome were apparent even on day-of-admission, prior to the development of clinically defined AKI. Selected candidate biomarkers were validated in an independent larger cohort of ACLF patients. Our study provides insights into the dynamic changes that occur in the plasma proteome of ACLF patients prior to the onset of AKI, provides a potential predictive biomarker for AKI in ACLF and might have therapeutic potential as well based on its cellular targets. We report our findings here.

## STUDY DESIGN, PATIENT RECRUITMENT AND METHODOLOGY

### Study Design

An overall workflow of the study has been provided in Figure 1A. The study was divided into 2 phases- (A) Discovery phase and, (B) Validation phase (Figure 1A). The discovery phase aimed at a de novo identification of plasma proteins which were significantly altered (up- or down-regulated) in ACLF patients who did not have clinically defined AKI at admission but progressed to AKI within 10 days. Therefore, day-of-admission (Day 0) plasma samples were collected from ACLF patients without AKI at presentation (n=19). These patients were followed up over a 10 day period for the development of AKI as defined by the criteria below. At the end of 10 days, patients were classified as either those who progressed to AKI (ACLF progression to AKI; n=10) or those who did not progress to AKI (ACLF no-progression to AKI, n=9). The Day 0 plasma samples of these patients were then subjected to label free quantitative proteomics and differentially expressed proteins were identified. Candidate biomarker proteins were selected on the basis of their relative fold up regulation in ACLF patients who progressed to AKI. The second phase of the study was the validation phase, where the identified biomarker proteins were quantitatively measured using ELISA in a larger cohort of 93 ACLF patients (33 ACLF-AKI on presentation and 60 ACLF no-AKI) (Figre 1A). Among the 60 ACLF patients without AKI (ACLF no-AKI), 20 progressed to AKI within 10 days (progression to AKI) whereas 40 did not progress to AKI (No-progression to AKI). Day 0 plasma samples were collected from all the study participants which was used for biomarker validation by ELISA. Since this was a discovery-study, and the validation of identified biomarkers was carried out for the first time, a sample size determination was not carried out.

**Figure 1.**
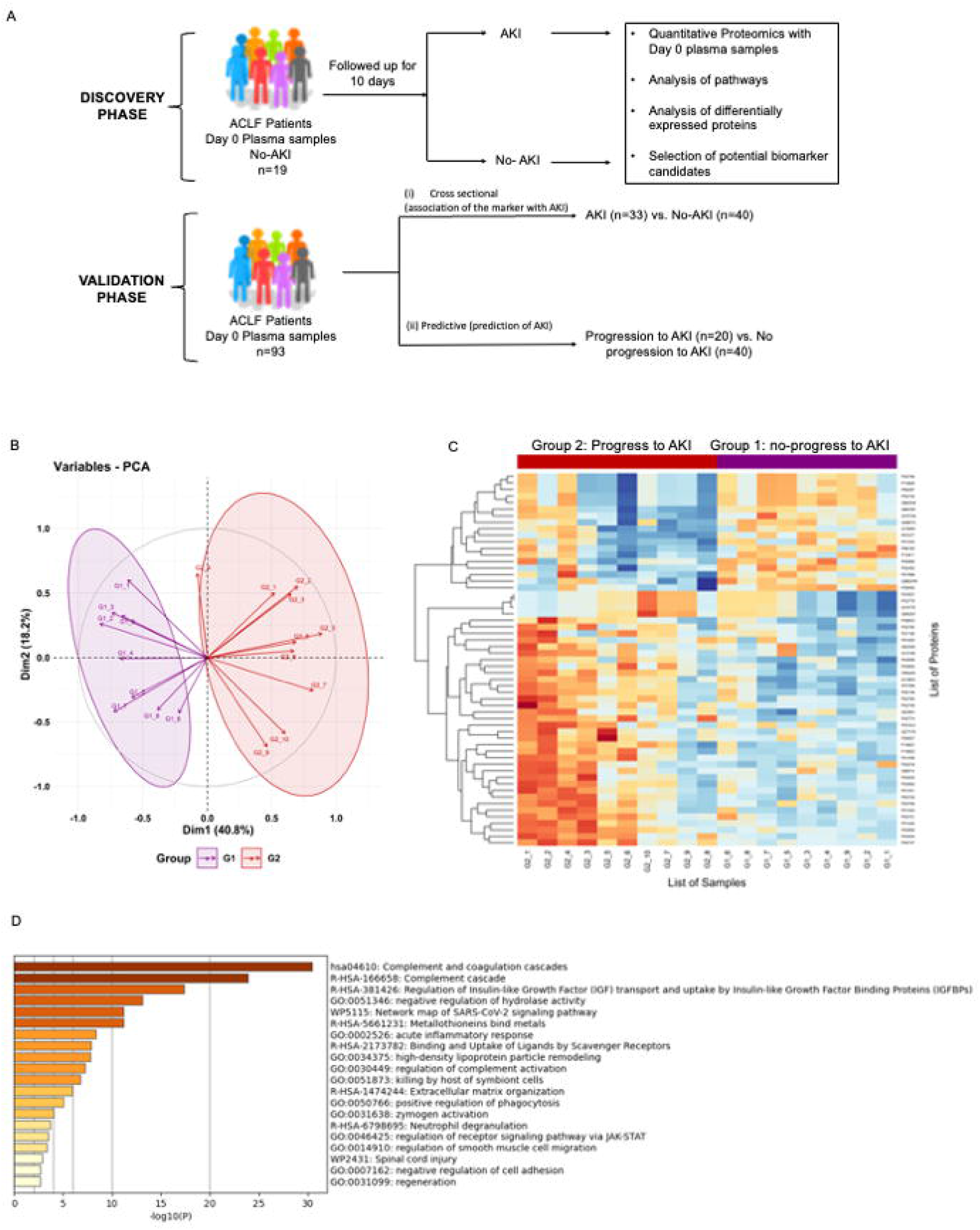
Proteomics Analysis of ACLF patients who progress to AKI vs ACLF patients who do not progress to AKI. (A)Study design of proteomics analysis. (B) PCA analysis clusters patient population into two distinct groups based on plasma proteome profiles, (C) Heatmap of protein expression in the two different groups (G1: ACLF patients who do not progress to AKI and G2: ACLF patients who progress to AKI) and (D) Pathways analysis using Metascape suggests an upregulation of coagulation, acute inflammatory pathways and metal binding proteins metallothioneins as major pathways upregulated in the plasma of ACLF patients who progress to AKI.

The patients gave written informed consent for participation in the study and the study was approved by the All India Institute of Medical Sciences, New Delhi ethics committee [Reference No. IEC/473/9/2016 and, IEC/369/7/2016], which is in agreement with the declaration of Helsinki^10^

All patients were managed as per standard uniform protocol described earlier from our center^11^.

### Recruitment of Patients

Consecutive patients diagnosed with acute-on-chronic liver failure (ACLF) and admitted in the Department of Gastroenterology, All India Institute of Medical Sciences, New Delhi, India were included. The patient recruitment period was between April 2017 and November 2021. A diseased control group (patients with compensated Cirrhosis, chronic liver disease (CLD)) who presented to the Gastroenterology OPD were included in the study to identify the baseline biomarker levels in patients with compensated cirrhosis of liver.

### Diagnostic and Exclusion Criteria

ACLF was diagnosed as per the EASL (European Association for the Study of Liver) criteria as an acute deterioration of pre-existing chronic liver disease, usually related to a precipitating event and associated with increased mortality at 3 months due to multisystem organ failure and ACLF grades were defined as per EASL-CLIF consortium criteria^12^. Compensated cirrhosis was diagnosed based on conventional clinical, biochemical, endoscopic and imaging as well liver histological criteriawere recruited from the Department of Gastroenterology. Such patients were within the age group□>□18 years and□<□75 years, treatment naïve, ambulatory, without any extra-hepatic complications, and no overt symptoms^13^.

AKI was defined as per the consensus recommendations of the International Club of Ascites-increase in sCr ≥0.3 mg/dL (≥26.5 mmol/L)from base line within 48 h; or a percentage increase sCr ≥50% from baseline which is known, or presumed, to have occurred within the prior 7 days^14^.

Exclusion criteria were hepatocellular carcinoma or portal vein thrombosis, age □<□18 years and□>□75 years, presence of prior renal, respiratory and/or cardiovascular disease.

### Plasma preparation for proteomics analysis

Whole blood was collected in Becton Dickinson (BD) vacutainer EDTA vials within 24 hours of admission for plasma preparation. Processing of whole blood was done within 2 hours of sample collection. Briefly, the blood vial was centrifuged at 400 x g for 10 minutes at room temperature two times. Platelet free plasma was separated and aliquoted in cryo-vials and stored at -80°Celsius until further use.

### Protein Isolation and sample preparation

Total protein concentration of each of the plasma samples were estimated using BCA kit (Puregene Cat no. GX6410AR). Plasma corresponding to 600 μg of protein were subjected to depletion using Thermo Scientific Top 12 Abundant protein depletion column.Depleted plasma samples were submitted for trypsin digestion (Trypsin Gold, Promega) and clean-up using C18 columns (NEST group microspin silica column). 50 μg of the Gn-HCL protein lysate were first reduced with 5 mM TCEP and further alkylated with 50 mM iodoacetamide. Alkylated proteins were further diluted using 50mM ammonium bicarbonate to bring final Gn-HCL concentration to 0.6M and then digested with trypsin (1:50, trypsin: lysate ratio, Promega) for 16 h at 37°C. The overnight digests were clarified with brief spin and the supernatant pH was adjusted around pH2 using 10% TFA.Digests were cleaned using a C18 silica cartridge to remove the salt and dried using a speed vac. The dried pellet was resuspended in buffer A (5% acetonitrile, 0.1% formic acid). The mass spectrometric analysis was carried out at vProteomics, New Delhi.

### Preparation of RNA, cDNA and PCR from Post Mortem Liver Tissue Biopsies

Tru-cut liver biopsies were retrieved from ACLF within 30 minutes of death with informed consent, and immediately stored in RNA Later in the AB2 ward of the Department of Gastroenterology, AIIMS New Delhi. The biopsied tissue were transferred to the Department of Biochemistry within 1-12 hours of collection and transferred to Trizol for long term storage, and stored at -80°C until further use. This part of the study received a separate ethical clearance from the Institute Ethics Committee (Ref. No.IEC/687/8/2019).

Prior to use, RNA was prepared from the tissue samples using a Qiagen RNeasy miniprep kit (Cat. No. 74106)as per the manufacturer’s instructions. RNA quantity was measured using a nanodrop machine and 500 ng of RNA per tissue sample was subjected to cDNA preparation using the Thermofisher verso cDNA preparation kit (Cat. No.AB1453B). Primers for MT1 A and MT2 were designed on the basis of a previously published study^36^. The published primers were subjected to a nucleotide BLAST analysis in order to ensure that they were specific to human MT genes as expected. The primer sequences used in this study were as follows:

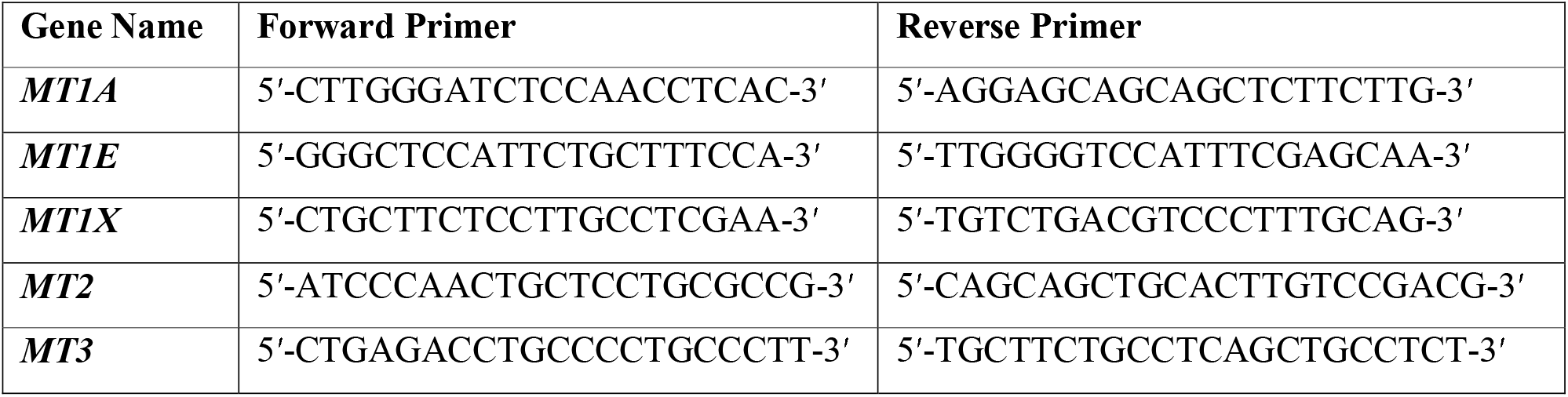

cDNA obtained from the post-mortem liver biopsies of deceased ACLF patients were subjected to Reverse Transcriptase-PCR analysis and the PCR products were analyzed on a 1% agarose gel along with molecular weight markers.

### Mass Spectrometric Analysis of Peptide Mixtures

Mass spectrometric analysis of the plasma samples was performed at vProteomics (New Delhi). The trypsin-digested sample (1 μg) was resolved on a 50-cm long PicoFrit column (360 μm outer diameter, 75 μm inner diameter &10 μm tip) filled with 2.0 m-C18 resin on nano1000 chromatography system (Proxeon, Thermo) attached to QExactive mass spectrometer. The peptides were eluted with a 5-15% gradient of the Buffer-B (95% acetonitrile, 0.1% formic acid) for 85 min, 15-40% gradient for 80min, followed by 95% gradient for 6 min at a flow rate of 300 nl/min for the total run time of 100 min. The QExactive spray voltage was set at 2.5 kV, S lens RF level at 50 and ITC heated capillary temperature at 275°C. The MS data were acquired in positive polarity using a data-dependent method choosing 10 most intense peaks with charge state +2 to+5, with exclude isotope option enabled and dynamic exclusion of time of 12 sec. The MS1 (mass range 150-2000 m/z) and MS2 scans were acquired in Orbitrap Mass analyzer with resolution of 70,000 and 17500 at m/z 200 respectively with lockmass (445.12003) option enabled. The MS1 or Full scan target was 1×10^6^ with a maximum fill time of 100 ms with mass range set to 350−1700. Target value for MS2 or fragment scans was set at 1×10^5^, and intensity threshold was set at 5×10^3^. Isolation window of parent ion of interest was set at 2 m/z. Normalized collision energy for Higher-energy collisional dissociation (HCD) was set at 27. Peptide match option was set to preferred mode along with activation of isotope exclusion option.

### Data Processing and Differential Proteome Analysis

All samples were processed and the RAW files generated were analyzed with Proteome Discoverer (v2.4) against the Uniprot reference proteome database as provided. For Sequest and Amanda search, the precursor and fragment mass tolerances were set at 10 ppm and 0.5 Da, respectively. The protease used to generate peptides, i.e., enzyme specificity was set for trypsin/P (cleavage at the C terminus of “K/R: unless followed by “P”) along with maximum missed cleavages value of two. Carbamidomethyl on cysteine as fixed modification and oxidation of methionine and N-terminal acetylation were considered as Variable modifications for database search. Both peptide spectrum match and protein false discovery rate were set to 0.01 FDR.

Abundance values of each sample were used for differential statistical analysis. Protein abundance values were filtered on the basis of valid values. Filtered values were Log2 transformed followed by Z-score standardization. Student T-Test wasused as the numbers of groups according to the study and statistical significance was considered for p-values ≤0.05. A p-value ≤ 0.05 was considered significant. Z score abundance values of the Significant proteins were then used for bioinformatics data visualization using in-house R Programming Scripts (vProteomics, New Delhi). Box and whisker plot were drawn for log2 transformed and Z-score standardized abundance values.

HeatMapfor differentially expressed proteins was drawn using R programming. Group legends were used to categorize samples (reference G1 and test G2). Principle components analysis (PCA)was performed to see the dimensional spread of data variability. A Biplot (using PC1 and PC2) was used to visualize the spread of samples.

Pathways analysis were carried out using Metascape and information on protein expression were retrieved from human protein atlas (proteinatlas.org)^15-17^.

### ICP-MS for Elemental Analysis

Collected plasma samples were diluted in acid digestion matrix containing ultra-high pure n-butanol, EDTA, hydrogen peroxide, HNO_3_ and triton-x solution. The blood samples were diluted 100 times (specimen:digestion matrix ratio was 1:99). The specimens were allowed to be digested in the digestion matrix for 1 hour. After matrix digestion, specimens were vortexed for 5 minutes. The matrix was prepared using supra-pure metal free double distilled water.The same digestion matrix was used for calibration standard preparation. The multi calibration standards containing As, Cu, Mn, Se, Cd, Pb and Hg (Agilent Technologies, USA) were used to prepare multiple calibration matrix having different elemental concentrations (blank matrix, 0.1 ppb, 0.5 ppb, 1 ppb, 5 ppb, 10 ppb and 50 ppb). After warming-up and start-up sequence calibration, we calibrated the analytical batches using these standards.The internal standard (Agilent Technologies, USA) containing Lithium, Bismuth, Germanium, Scandium and Indium were diluted in the same matrix in which standard and samples were prepared. The internal standard was run with calibration standards and samples were analysed for the standard stability and accuracy of the concentration measurement.The certified standards of blood (Recipe, Germany) were used for quality control (QC). QC samples were prepared in the same matrix used for standard preparation and were analysed after every 6 analytical runs. The working standards were freshly prepared before analysis to avoid any interference and contamination. A standard analytical run included three replicates for each specimen and for each replicate the machine recorded 50 readings. We compared the quality of the analysis and concentration measurements with the internationally certified blood standards containing As, Cu, Mn, Se, Cd, Pb and Hg.

### Validation of Selected Differentially Expressed Plasma Proteins

Metallothionein (MT) proteins were selected for further validation in ACLF and CLD patient plasma through ELISA. Protocols were followed as per manufacturer’s suggestions for both the Human Metallothionein (MT) ELISA kit (CUSABIO, Cat no. CSB-E09060h). All plasma samples were used at a dilution of 1:200. ELISA results were read on a MicroTek multimode reader. ELISA results were analyzed on MS Excel and statistical analysis was carried out using GraphPad Prism9.0.

### Statistical Analysis

Data was analysed by statistical software Stata 14.0. Variables were checked for approximate normal distribution. Quantitative data were expressed as Mean±SD and Median (Interquartile range). Categorical data was expressed as frequency (%). Chi-squared test/Fisher’s exact test was used to compare categorical variables between the groups. Indepednt t test was used to compare quantitative variable btween the groups. Those variable that did not follow normal distribution were compared by ranksum test. One way ANOVA /Kruskal Wallis test, followed by Bonferroni/Dunn test correction used to compare quantitative variables among different list of variables. ROC analysis was carried out to find the sensitivity and specificity of the MT (ng/ml) corresponding to optimum cut-off. Logistic regression used to estimate the odds of AKI adjusted for covariates. Time to event analysis (Kaplan-Meier curve) used to compare probability of an event (no-progression to AKI) at a certain time interval. All statistical tests used were two-sided and p-value of less than 0.05 was used to indicate a statistically significant difference.

## RESULTS

### Baseline Characteristics of Patients

93 patients were included in the study. The study had 2 phases- (i) The discovery phase and (ii) The validation phase (Figure 1A).

#### (i) The discovery phase

Plasma proteome analysis in order to identify differentially expressed proteins in ACLF patients who progressed to AKI vs those who did not. This part of the study included the following patient groups:

The study groups included in the proteomics analysis were- (i) ACLF patients who did not progress to AKI (Group1; n□=□9) and, (ii) ACLF patients who progressed to AKI (Group 2; n=10) (Table 1). The mean ages in Group1 and Group2 were 43± 4 years and 47 ± 4 years respectively. There were no differences in the laboratory parameters between ACLF patients who developed vs did not develop AKI (Table 1).

**Table 1.** Baseline characteristics of ACLF samples subjected to proteomic analysis. Group 1 (G1; n=9) represents ACLF patients who do not progress to AKI and Group 2 (G2; n=10) represents ACLF patients who progress to AKI.

|  | **G1 (n=9)** | **G2 (n=10)** | **p-value** |
| --- | --- | --- | --- |
| **Age (Mean, years ± SD)** | 43 ± 4 | 47 ± 4 | 0.42 |
| **Males (%)** | 66.60 | 70 | 0.999 |
| **Hb (g/dL)** | 9.189 ± 0.9710 | 7.980 ± 0.8012 | 0.35 |
| **TLC (mEq/L)** | 8300 (6640-10650) | 13545 (5125-18825) | 0.31 |
| **Platelet (x1000)** | 82 (43.5-153) | 40 (24.25-66.5) | 0.11 |
| **INR** | 1.93 (1.75-3.12) | 2.95 (2.06-3.77) | 0.08 |
| **Urea (mg/dL)** | 53 (20-68) | 78.5 (50-121) | 0.08 |
| **Creatinine (mg/dL)** | 1.133 ± 0.1291 | 1.200 ± 0.1660 | 0.76 |
| **Albumin (g/dL)** | 2.478 ± 0.1824 | 2.910 ± 0.2345 | 0.17 |
| **Sodium (mEq/L)** | 142.2 ± 2.332 | 140.3 ± 4.033 | 0.70 |
| **Potassium (mEq/L)** | 4.198 ± 0.3004 | 4.300 ± 0.3756 | 0.83 |
| **Bilirubin (mg/dL)** | 15.10 ± 3.096 | 18.57 ± 3.456 | 0.46 |
| **AST (IU/L)** | 101 (75-147.5) | 136 (75.5-284) | 0.40 |
| **ALT (IU/L)** | 47 (22.5-80.5) | 42 (32.25-58.5) | 0.94 |
| **SAP (IU/L)** | 199 (161.5-451) | 243 (141-337.5) | 0.94 |
| **ACLF Grades** |  |  |  |
| **Grade 2** | 6 | 1 |  |
| **Grade 3** | 3 | 9 |  |

#### (ii) The validation phase

ELISA based validation of identified biomarkers in the discovery phase. This included the following study groups:

#### (A) ACLF AKI vs ACLF no-AKI

For the validation studies, n=93 ACLF patients were recruited and n=36 CLD patients were recruited (Tables 2 and 3). Plasma samples were collected on the day of admission from all study participants and prepared within 2 hours of collection. ACLF patients were further classified in two different ways- (i) ACLF AKI and ACLF no-AKI, based on the presence or absence of AKI on the day of admission (Table 2) or (ii) into ACLF-progression to AKI and ACLF no progression to AKI. The latter classification was done with only those patients who were ACLF no-AKI on the day-of-admission and were followed up for the appearance of AKI within 7-10 days of admission (Table 3). Baseline comparison between ACLF AKI, ACLF no-AKI and CLD showed that the mean age of patients in all the three groups were comparable (mean ages were 43 ± 12 years, 43 ± 12 years and 41 ± 12 years respectively; Table 2). A majority of the patients in all three groups were males. Urea and creatinine levels were significantly higher in ACLF AKI vs ACLF no-AKI, as expected (median creatinine levels were 2.7 mg/dL and 1.0 mg/dLrespectively, p-value ≤0.001; median urea levels were106 mg/dL and 48 mg/dL respectively, p-value ≤0.001; Table 2). TLC values were also higher in ACLF AKI vs ACLF No-AKI (median values 15330/mm^3^ and 9330/mm^3,^ respectively, p-value ≤0.001; Table 2).

**Table 2:** Baseline characteristics across groups. ACLF no-AKI: ACLF patients without AKI; ACLF-AKI: ACLF patients with AKI; CLD: Chronic Liver Disease (various etiology). All values are measured on the day of diagnosis (Day 0). All values are Mean ± SD or Median (Range). ^*^ denotes significant differences in comparisons between CLD vs ACLF no-AKI; and CLD vs AKI; ^$^ denotes significant differences in comparisons between ACLF no-AKI vs ACLF AKI.

|  | CLD (n=35) | ACLF |  | ANOVA |
| --- | --- | --- | --- | --- |
|  |  | No-AKI (n=60) | AKI (n=33) | P value |
| **Age, years, Mean ± SD** | 41 ± 12 | 43 ± 12 | 43 ± 12 | 0.701 |
| **Males %** | 30 (85.7%) | 46 (76.7%) | 28 (84.85%) | 0.457 |
| **Hemoglobin (g/dL)** | 10.7 ± 2.7 | 8.4 ± 2.1* | 7.5 ± 1.6* | 0.013 |
| **Na (mEq/L)** | 138.9 ± 4.9 | 139.6 ± 9.1 | 137.4 ± 11.6 | 0.522 |
| **Albumin (g/dL)** | 3.5 ± 0.9 | 2.8 ± 0.9* | 2.6 ± 0.4* | <0.001 |
| **TLC (x103 per mm3)** | 6630 (5260-9400) | 9330 (6280-12585)* | 15330 (10470-25630)*\$ | <0.001 |
| **Platelet (x103 per mm3)** | 110 (92-152) | 71 (50-103.5)* | 80 (57-110)* | 0.001 |
| **Urea (mg/dL)** | 22 (17-30.5) | 48 (24-69)* | 106 (78.2-167)*\$ | 0.0001 |
| **Creatinine (mg/dL)** | 0.8 (0.7-0.9) | 1.0 (0.7-1.4) | 2.7 (2.2-4.6)*\$ | 0.0001 |
| **Potassium (mEq/L)** | 4.2 ± 0.5 | 4.5 ± 2.5 | 4.7 ± 1.2 | 0.443 |
| **Bilirubin (mg/dL)** | 2 (0.9-4) | 11 (4.5-22.1)* | 8 (4.1-23.5)* | 0.0001 |
| **AST (IU/L)** | 46 (37-72) | 96 (56.5-178.5)* | 82 (59-206)* | 0.0001 |
| **ALT (IU/L)** | 29 (22-42) | 39 (25.5-62)* | 42 (29-79)* | 0.009 |
| **SAP (IU/L)** | 297 (188-410) | 199 (131-277.5)* | 185 (158-221)* | 0.018 |
| **MELD** | - | 25.2 ± 8.0 | 36.7 ± 8.0 | <0.001 |
| **MELD Na** | - | 24.4 ± 9.3 | 36.0 ± 10.1 | <0.001 |
| **CLIF-OF** | - | 10.1 ± 3.1 | 11.8 ± 2.9 | 0.032 |
| **CTP** | - | 12.1 ± 2.0 | 12.1 ± 2.5 | 0.944 |
| **MT (ng/ml)** | 4.2 (3.0-8.1) | 4.6 (1.8-10.5) | 13.6 (5.9-25.5)*\$ | 0.0001 |
| **Infection (% of patients)** | - | 36 (60%) | 27 (81.8%) |  |
| **In-hospital stay non-survivor** |  | 28 (46.6%) | 26 (78.7%) | 0.006 |
| **28-day non-survivor** |  | 31 (51.7%) | 28 (84.8%) | 0.004 |
| **90 days non survivor** |  | 35 (58.3%) | 28 (84.8%) | 0.128 |

**Table 3:** Baseline characteristics of ACLF patients who progress to AKI and who ACLF patients who do not progress to AKI. All values are measured on the day of diagnosis (Day 0). All values are Mean ± SD. * Indicates those parameters which are significantly different in groups 1 and 2 (* p <0.05; ** p<0.01, *** p<0.001)

|  | **Group 1** | **Group 2** |  |
| --- | --- | --- | --- |
|  | **No-progression to AKI (n=40)** | **Progression to AKI (n=20)** | **P value** |
| **Age, years, Mean ± SD** | 42 ± 2 | 45 ± 10 | 0.356 |
| **Males %** | 32 (76.19%) | 17 (80.95%) | 0.666 |
| **Hemoglobin (g/dL)** | 8.6 ± 2.14 | 7.79 ± 1.95 | 0.135 |
| **Na (mEq/L)** | 138.69 ± 7.95 | 139.85 ± 11.73 | 0.461 |
| **Albumin (g/dL)** | 2.83 ± 0.96 | 2.75 ± 0.70 | 0.794 |
| **TLC (x103 per mm3)** | 9000 (5500-12100) | 9630 (6460-19900) | 0.384 |
| **Platelet (x103 per mm3)\*\*** | 89 (60-120) | 50 (39-76) | 0.006 |
| **Urea (mg/dL)\*\*\*** | 31.5 (21-54) | 69 (51-110) | 0.0008 |
| **Creatinine (mg/dL)\*** | 0.8 (0.7-1.3) | 1.3 (0.8-1.7) | 0.026 |
| **Potassium (mEq/L)** | 4.15 (3.7-4.6) | 4.35 (3.6-4.7) | 0.131 |
| **Bilirubin (mg/dL)** | 8.75 (4.37-22.5) | 12.7 (4.9-16.9) | 0.500 |
| **AST (IU/L)\*** | 80 (49-146) | 131 (72-223) | 0.024 |
| **ALT (IU/L)** | 37.5 (24-57) | 45 (33-66) | 0.108 |
| **SAP (IU/L)** | 195 (115-280) | 210 (162-275) | 0.782 |
| **MT (ng/ml)\*\*\*** | 3.49 (1.68-5.58) | 10.18 (6.15-11.98) | 0.001 |
| **In-hospital stay non-survivor** | 13 (32.5%) | 15 (75.0%) | 0.004 |
| **28-day non-survivor** | 16 (40.0%) | 15 (75.0%) | 0.028 |
| **90 days non survivor** | 19 (47.5%) | 16 (80.0%) | 0.124 |

#### (B) ACLF progression to AKI vs ACLF no-progression to AKI

All the ACLF no-AKI patients were followed up for 10 days to evaluate the development of AKI and were retrospectively classified into ACLF no-progression to AKI (Group1) and ACLF progression to AKI (Group 2). Platelet counts (89 ×10^3^and 50 ×10^3^ per mm^3,^ p-value ≤0.01), urea levels (31.5 mg/dL and 69 mg/ dL, p-value≤0.001)and creatinine levels (0.8 and 1.3 mg/dL, p-value≤0.05) were found to be significantly different in ACLF progression to AKI vs ACLF no progression to AKI respectively (Table 3).

### Plasma Proteomics of ACLF-Progression to AKI patients vs ACLF No progression to AKI patients

The major aim of the proteomics experiment was to discover plasma proteome changes that occur prior to the onset of clinically defined AKI and can act as predictive biomarkers of AKI in ACLF. Therefore, from the cohort of ACLF patients who were enrolled into the study, we retrospectively selected 19 ACLF patients with no-AKI on the day of admission; as per the AKIN criteria. Of these 19 patients (ACLF No-AKI), 9 patients did not develop AKI over the next 28 days of follow up (Group 1-ACLF No progression to AKI) and 10 patients developed AKI within the next 7 days (Group 2-ACLF progression to AKI). The day of admission (Day 0) plasma of these patients were collected and subjected to label free quantitative mass spectrometry as described in methods (Figure 1A). The proteome profiles of the two groups were found to be significantly different from each other (Figure 1B, C). The proteome profiles of Groups 1 and 2 were able to separately cluster the two groups in PCA analysis (Figure 1B). About 56 differentially expressed proteins were found in Group 2 as compared to Group 1 (ACLF-Progression to AKI) as compared to Group 1 (No-progression to AKI) (Figure 1C, Table 4). Pathways analysis using Metascape revealed signatures of activation of the complement system and coagulation cascade (circulatory failure), activation of metal binding pathway (novel pathway to be detected) and acute inflammatory response (Figure 1D). Among the highest upregulated plasma proteins, metallothioneins formed a prominent group; metallothionein isoforms MT1E, MT1M, MT1X and MT2 were upregulated in those ACLF patients who progressed to AKI (Table 3, Table 4; Figure 1D).

**Table 4.**
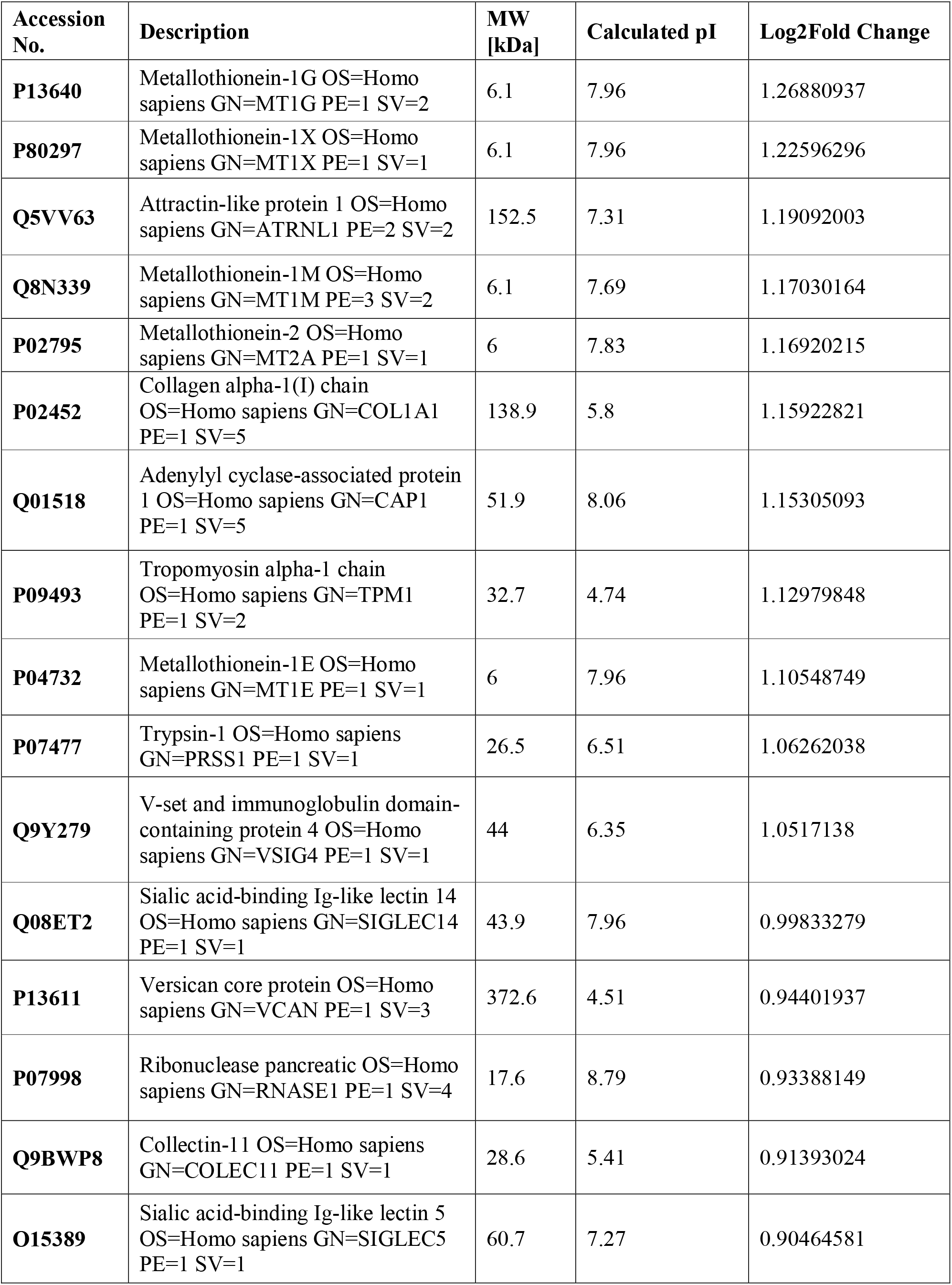
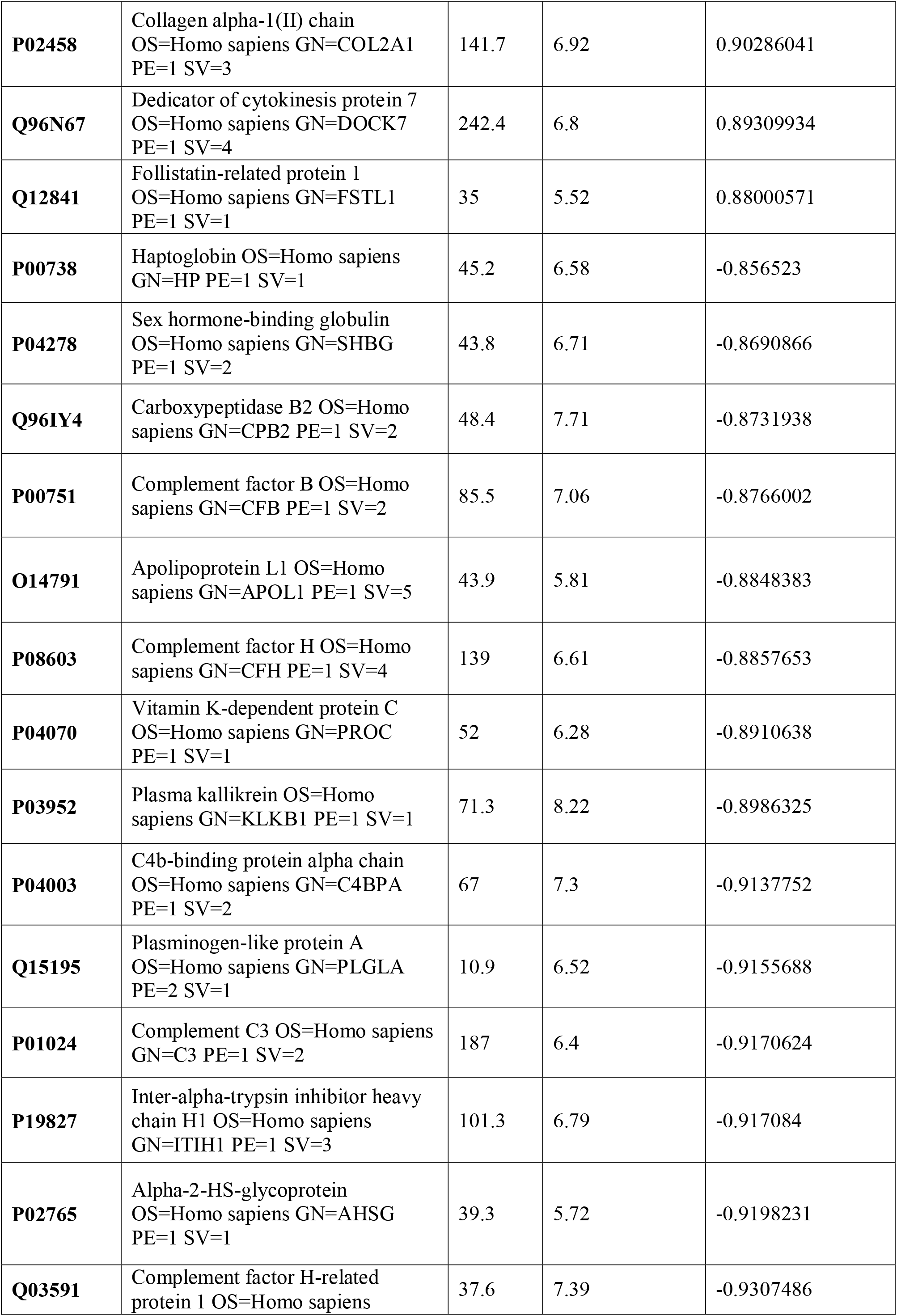
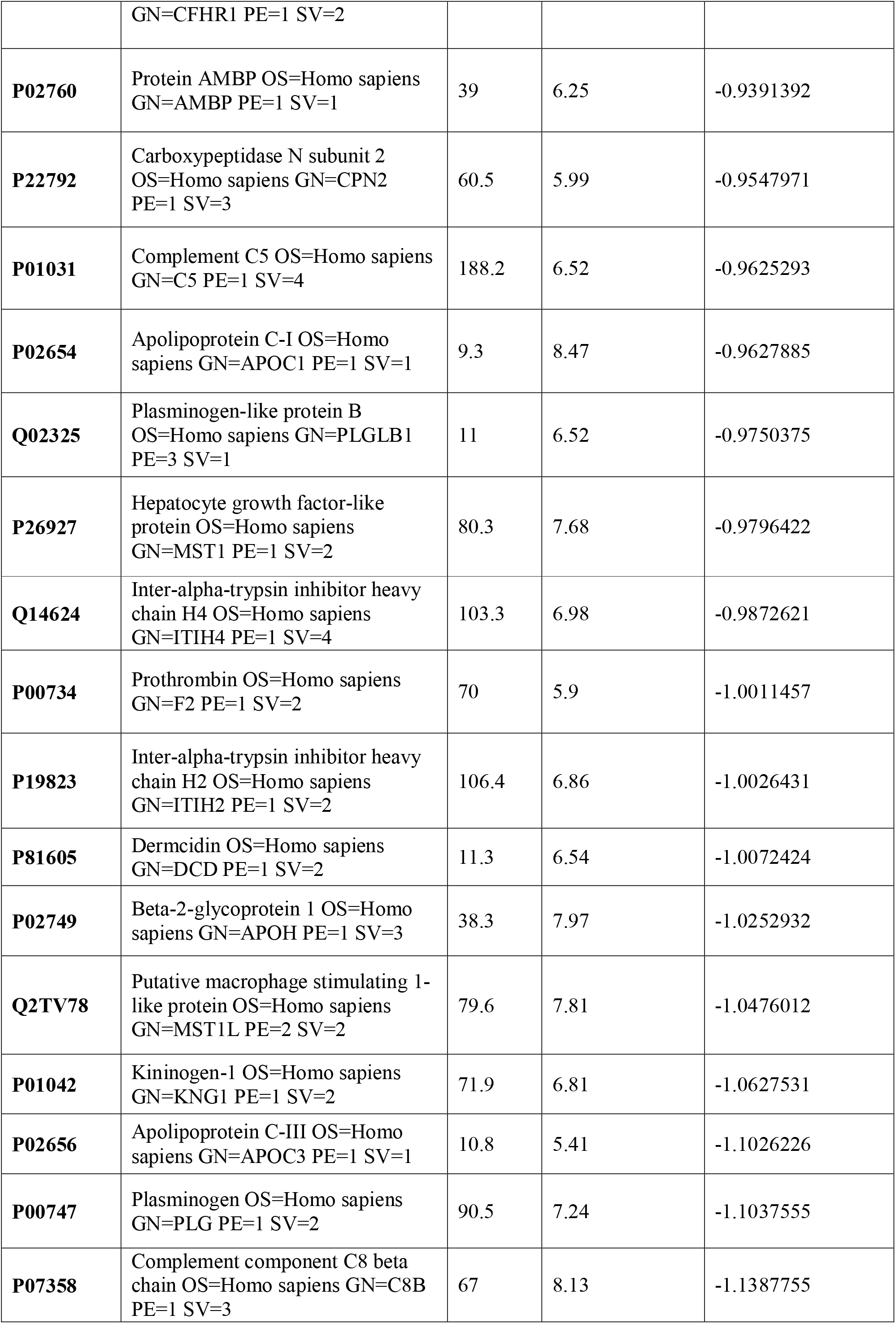
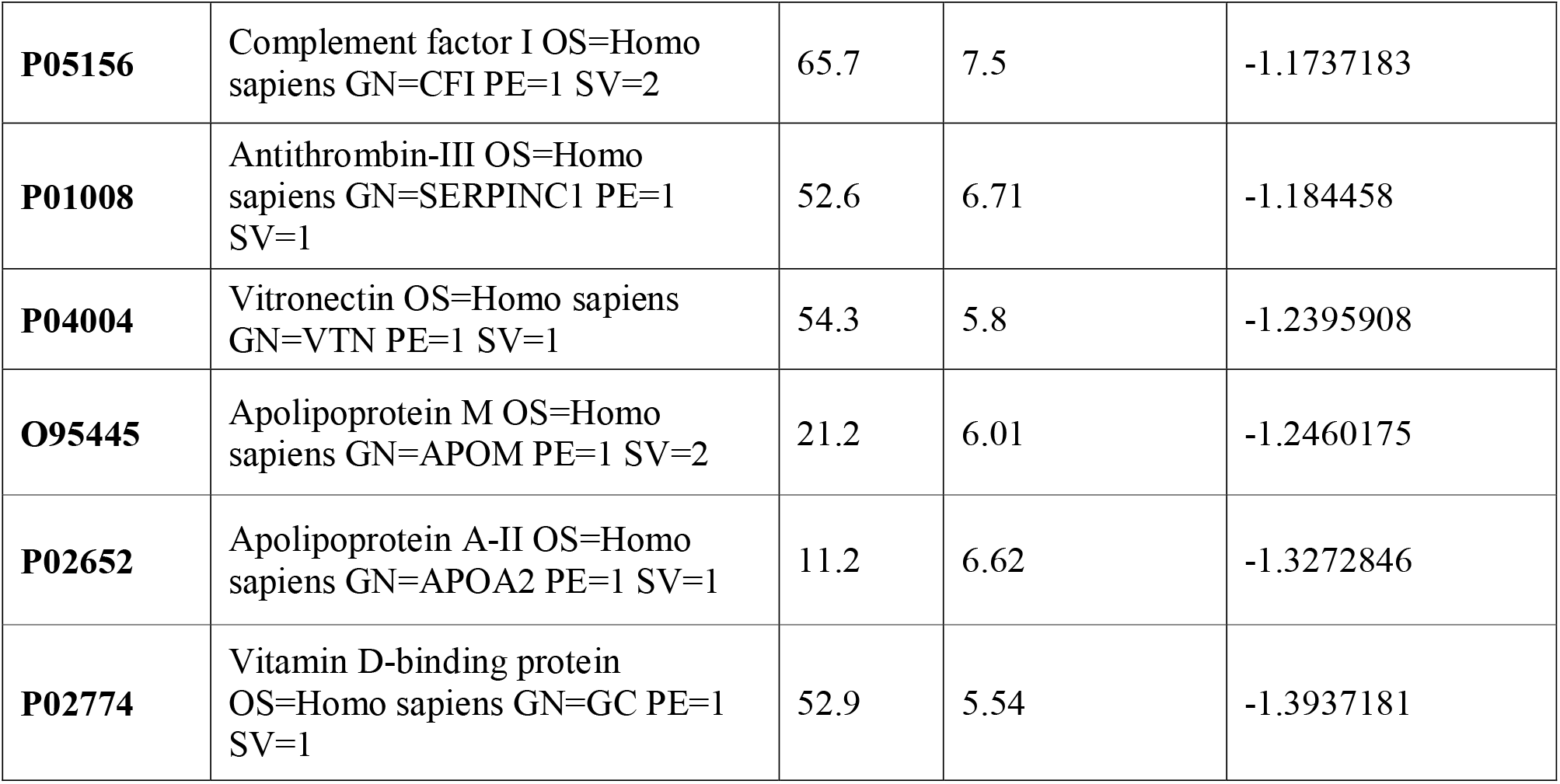
Differentially expressed proteins in Group 2 (ACLF progression to AKI; n=10) vs Group 1 (ACLF no-progression to AKI; n=9).

Cell type enrichment analysis revealed the presence of liver-enriched proteomic signature in the plasma proteome of ACLF patients who progressed to AKI (Supplementary Figure 1).Protein expression data from the human protein atlas (proteinatlas.org) showed that MT proteins are liver enriched genes (Figure 4B; proteinatlas.org). In addition, the plasma proteomic signature for ACLF patients who progress to AKI revealed a close resemblance with DisGeet signatures associated with acute kidney injury (AKI, Supplementary Figure 2).

### Plasma Metallothionein levels in ACLF patients who progress to AKI

ELISA based quantification confirmed that in ACLF-AKI, plasma MT levels (MT1 + MT2) were higher than in CLD patients (Figure 2A). Further, plasma MT levels were higher in ACLF -AKI vs ACLF No-AKI (p-value≤ 0.0001) and in ACLF progression to AKI vs ACLF no progression to AKI (p-value≤ 0.001; Figures 2A, B). The area under the receiver operating characteristics curves (AUROC) for ACLF-AKI vs ACLF no-AKI was 0.786(95% C.I. 0.66 - 0.85; p-value≤0.001) and for ACLF progression to AKI vs ACLF no progression to AKI was 0.7888 (95% C.I. 0.64-0.91, p-value≤0.001) (Figures 2C, D, Table 5).

**Figure 2.**
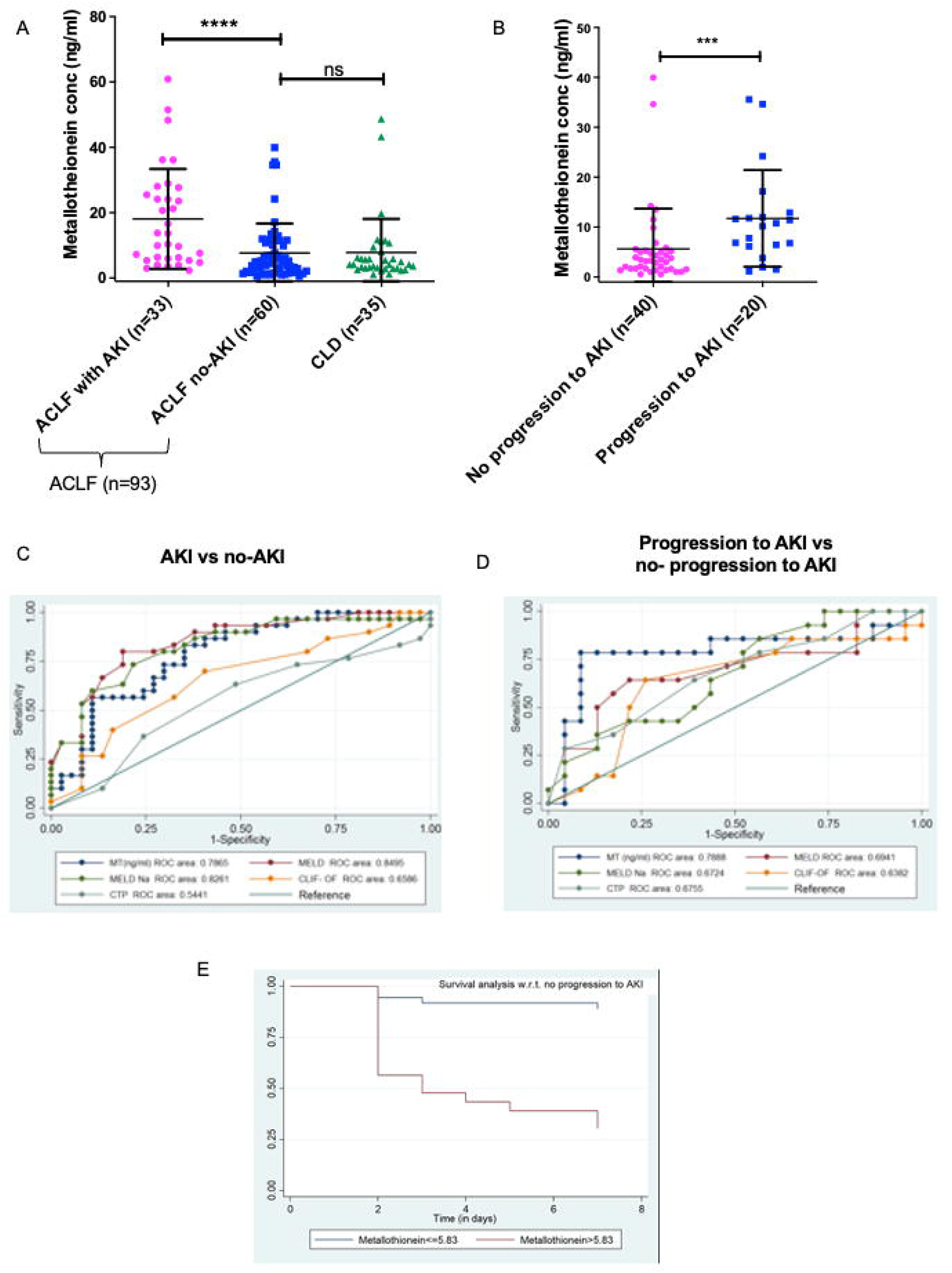
ELISA validation of plasma metallothionein (MT) levels in day-of-admission plasma from ACLF and CLD patients. (A,C)MT levels are significantly higher in ACLF-AKI patient plasma as compared to ACLF no-AKI or CLD with an AUROC of ∼ 0.79 (p-value ≤ 0.0001); (B, D) MT levels are significantly higher in plasma of ACLF patients who progress to AKI vs those who do not with an AUROC of ∼0.79(*** p-value ≤ 0.001; **** p-value ≤ 0.0001). (E) Kaplan-Meier analysis for progression to AKI for ACLF patients with plasma MT concentrations > 5.83 ng/ mL (p-value <0.0001).

**Table 5.** ROC analysis results of (a) ACLF AKI vs ACLF no-AKI and, (b) Progression to AKI vs No-progression to AKI

| Group |  | Cut off | Sensitivity | Specificity | AUROC (95% CI) |
| --- | --- | --- | --- | --- | --- |
| **ACLF AKI vs ACLF no-AKI** | MT (ng/ml) | $\leq 5.40$ | 78.79% | 58% | 0.7865 (0.66-0.85) |
| **No-progression to AKI vs Progression to AKI** | MT (ng/ml) | $\leq 5.83$ | 80.00% | 80.00% | 0.7888 (0.64-0.91) |

A discriminatory cut-off of 5.4 ng/ mL (78.8% sensitivity and 58% specificity) was determined for ACLF AKI vs ACLF no-AKI; and a discriminatory cutoff of 5.83 ng/ mL (80% specificity and 80% sensitivity) was determined for ACLF progression to AKI vs no-progression to AKI (Table 5). Univariate and multivariate analysis (adjusted for age, gender, ACLF Grade and infection) were carried out in order to calculate the odds ratio (OR) of having AKI and of progression to AKI with these cutoffs. The OR of having AKI, based on a cut-off of 5.4 ng/ mL was 4.38 (univariate; 95% C.I. 1.7-11.3) and (adjusted; C.I. 5.3 (1.3-21.3) (Table 6). The OR of ACLF patients progressing to AKI within 10 days of admission, based on a cut-off of 5.83 ng/ mL was 18.9 (univariate; C.I. 4.8-73.9) and 76.0 (adjusted, C.I. 1.67-3459) (Table 6).

**Table 6.** Multiple logistic regression analysis to discriminate between (a) AKI vs no AKI and (b) progression of AKI in ACLF patients without AKI, based MT levels (ng/ml) cut-off.

| Variable | Control | Case | P value | Univariate odds ratio (95%CI) | Adjusted odds ratio* (95%CI) |
| --- | --- | --- | --- | --- | --- |
| **MT ng/ml** |  |  | 0.002 | 4.38 (1.7-11.3) | 5.3 (1.3-21.3) |
| **<5.40** | 35 | 8 |  |  |  |
| **>5.40** | 25 | 25 |  |  |  |

\*Adjusted for age, gender, ACLF Grade, infection.

**b) No progression to AKI and progression to AKI**
| Variable | Control | Case | P value | Univariate odds ratio (95%CI) | Adjusted odds ratio (95%CI) |
| --- | --- | --- | --- | --- | --- |
| **MT ng/ml** |  |  | <0.001 | 18.9 (4.8-73.9) | 76.0 (1.67-3459) |
| **<5.83** | 31 | 4 |  |  |  |
| **>5.83** | 9 | 16 |  |  |  |

\*Adjusted for age, gender, ACLF Grade, infection.

A Kaplan-Meier survival analysis was carried out for metallothionein levels (MT) in 20 ACLF patients who progress to AKI within a follow up period of 7 daysfrom admission (Table 7). The cut off of 5.83 ng/ml was obtained from ROC analysis for ACLF patients who progress to AKI v/s patients with no progression to AKI. The survivor function (no-progression to AKI)was more likely for ACLF patients with MT levels < 5.83 ng/ml, whereas ACLF patients with MT levels >5.83 had a higher chance of developing AKI by day 7 (p-value <0.0001) (Table 7).

**Table 7.**
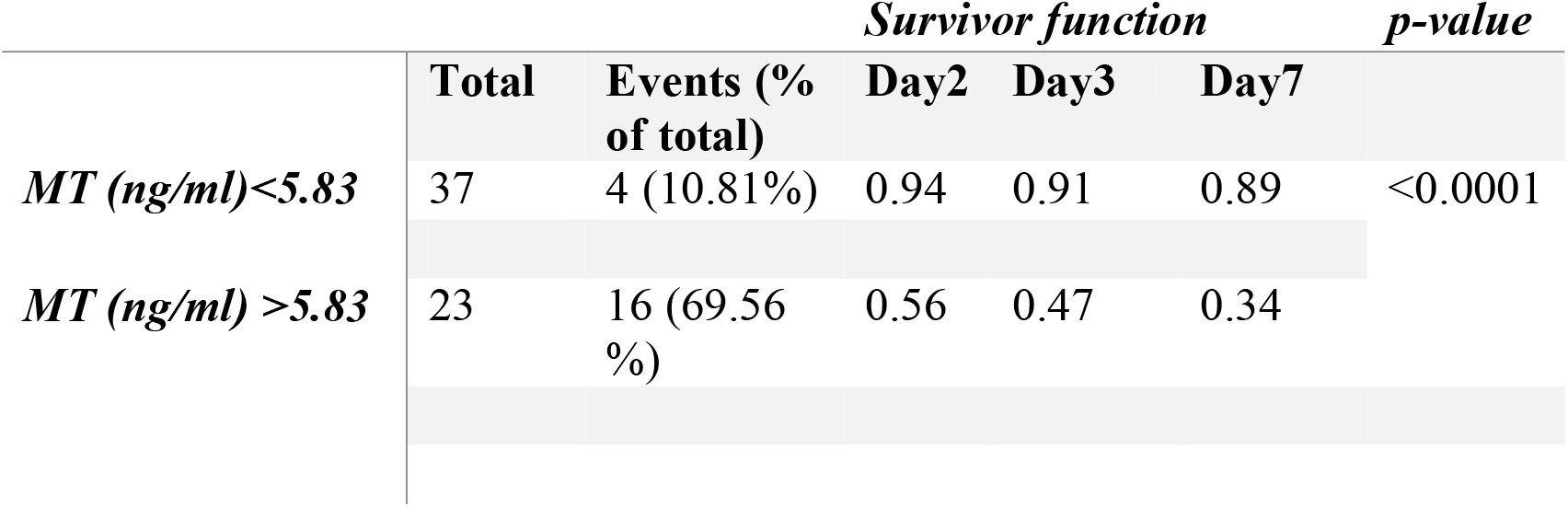
Kaplan Meier analysis for metallothionein levels (MT) (cut-off 5.83 ng/ mL) in 20 ACLF patients who progress to AKI within a follow up period of 7 days. The survivor function was no-progression to AKI.

In order to evaluate whether the induction of MT was due to an excess of metal ions, plasma elemental analysis was carried out by ICP-MS, as described in the methods. A small proportion of ACLF AKI patients showed elevated Ni levels in the plasma as compared to ACLF no-AKI and CLD (p-value= 0.06; Figure 3A). None of the other metals estimated showed significant differences between ACLF AKI and ACLF no-AKI or in ACLF vs CLD (Figures 3B-D).

**Figure 3.**
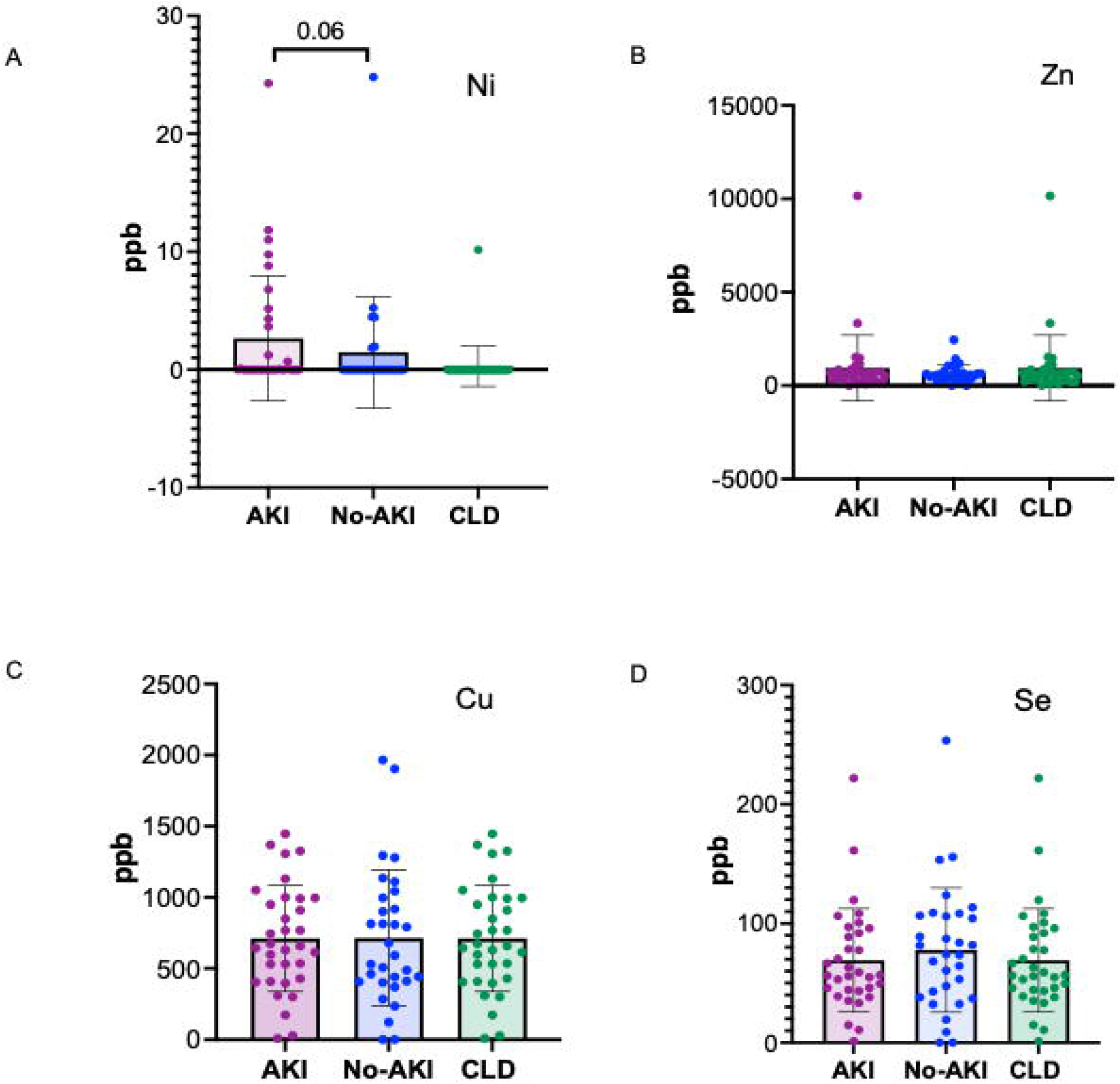
Plasma elemental analysis by ICP-MS for (A) Nickel, (B) Zn, (C) Cu and (D) Se. The levels of the metal ions are not significantly different in ACLF patients with and without AKI showing that heavy metal toxicity is not the cause for MT induction in ACLF. A fraction of ACLF AKI patients had elevated levels of Ni (A).

MT2 structure (downloaded from PDB) has 4 cysteine residues that coordinate with metal ions (https://www.rcsb.org/structure/1MHU; Figure 4A)^18^. The tissue protein expression graphs for all the isoforms of human MT were downloaded from human protein atlas (proteinatlas.org) and all MT were found to be liver enriched, although many were also expressed by other tissues (representative graphs Supplementary Figure 3, Figure 4B, proteinatlas.org)^16-17^. The protein atlas derived expression data for MT receptor megalin showed enrichment of megalin in the human proximal convoluted tubule epithelial cells (RPTECS) (Figure 4B, inset) ^16-17^.In order to examine if the ACLF liver is a potential source of MT, we carried out PCR analysis for MT1 and MT2 genes with cDNA derived from post-mortem liver biopsy from n= 7 ACLF patients (Supplementary Table 1). Robust expression of both MT1 and MT2 genes in the ACLF liver was observed (Figure 4C). The scavenger receptor megalin binds several plasma proteins as ligands such as albumin. In our study, serum albumin (Alb) levels were found to be significantly lower in ACLF vs CLD (p-value ≤ 0.0001; Figure 4D). Ratio of MT to Alb concentrations were calculated. MT/Alb was found to be significantly higher in ACLF vs CLD (p-value ≤0.01, Figure 4E), in ACLF-AKI vs ACLF no-AKI; (p-value ≤ 0.0001; Figure 4F) as well as in ACLF progression to AKI vs no progression to AKI, p-value≤ 0.0001; Figure 4G).

**Figure 4.**
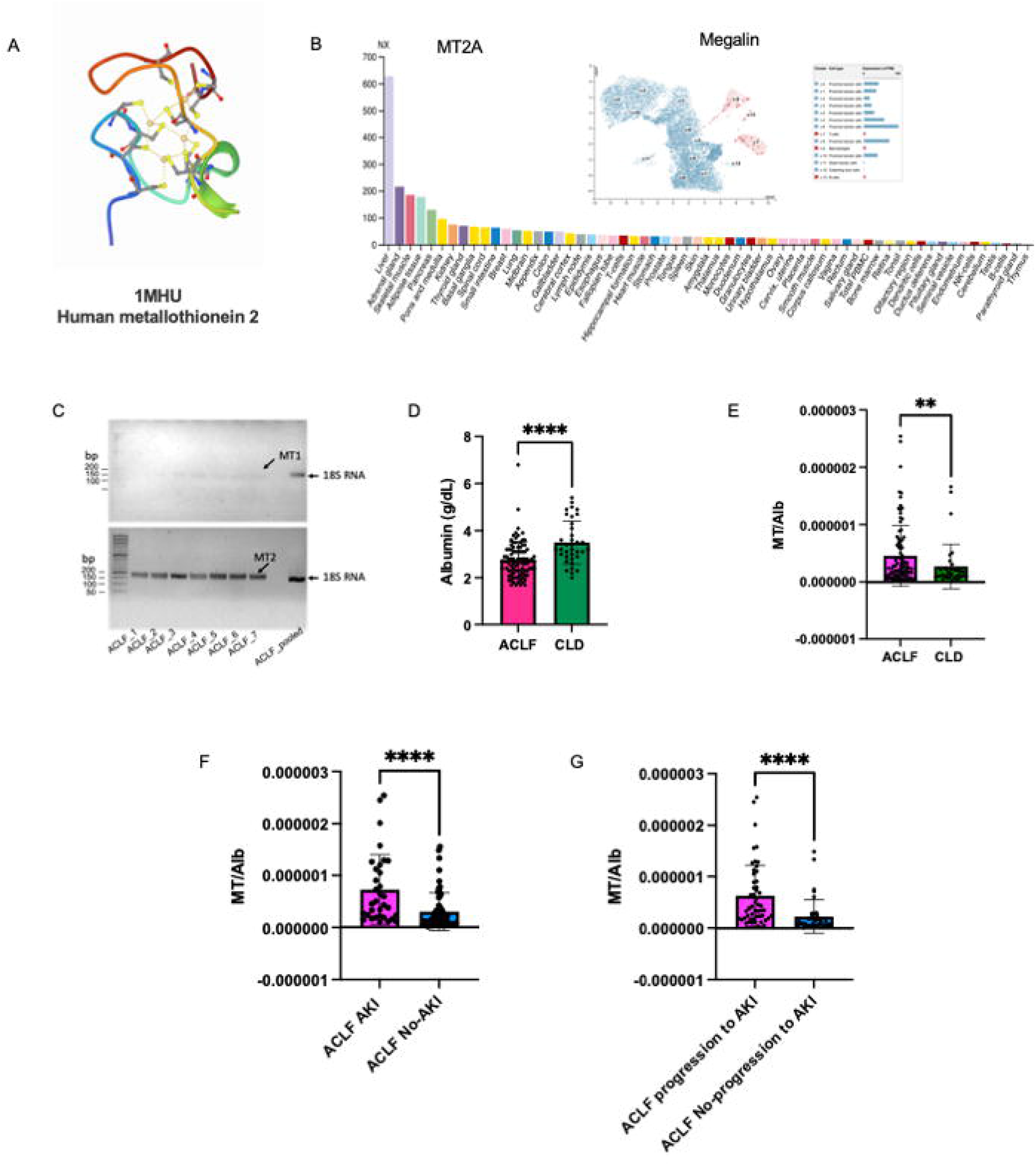
Metallothionein (MT) to Albumin (Alb) ratio in ACLF patients. (A) PDB structure of MT2A; (B) MT2A expression in various tissues shows that it is a liver enriched gene; (B, inset) Expression data for megalin, the scavenger receptor that binds MT, shows that it is expressed at high levels in renal PCT cells; (C) MT gene specific PCR from cDNA derived from post-mortem liver biopsy of ACLF patients shows robust expression of the MT2A gene and MT1A gene expression at a comparatively lower level; (D) Serum albumin (Alb) levels are significantly lower in ACLF-AKI patients compared to ACLF no-AKI; MT/Alb is significantly higher in (E) ACLF vs CLD; (F) ACLF AKI vs ACLF no-AKI and (G) ACLF patients who progress to AKI vs ACLF patients who do not progress to AKI. (**** p-value ≤ 0.0001).

## DISCUSSION

Acute kidney injury (AKI) is a defining feature of ACLF and significantly increases the risk of short-term mortality in ACLF patients^4^.However, there are no biomarkers for the prediction of AKI in ACLF patients, and the pathogenesis of AKI in ACLF is poorly characterized. The gold standard for renal function measurement is the quantitative estimation of serum urea and creatinine levels^37^. However these are neither very reliable nor sensitive biomarkers for renal function since their levels are influenced by multiple factors such as muscle mass, muscle injury, fluid therapy, and GFR^36^.Using quantitative proteomics approaches combined with ELISA based validation, our study demonstrated that the metallothionein (MT) family of proteins are highly upregulated on the dayofadmission in the plasma of ACLF patients at risk of developing AKI, prior to clinical onset of AKI. In addition, plasma MT concentrations were also significantly higher in ACLF patients with AKI on the day of admission (ACLF-AKI) as compared to those without AKI on the day of admission (ACLF no-AKI).Further, using AUROC, we were able to define a cut-off of 5.83 ng/ mL for plasma MT concentration, above which the odds of progression to AKI was 18.9 (C.I. 4.8-73.9) times that below the cut-off in univariate analyses and ∼70 (C.I. 1.67-3459) times in multivariate analysis adjusted for age, sex and the presence of sepsis, which is a common occurrence in ACLF. While the p-value of the OR calculations were significant (p≤ 0.001), the large C.I. suggest a high degree of variability in the measurements, i.e. a larger spread of data points. This is evident from Figures 2A and B wherein the ACLF patient samples exhibit a wide range of plasma MT values whereas CLD samples exhibit a relatively restricted range of values. Similarly, within ACLF, ACLF AKI and ACLF progression-to-AKI plasma MT concentrations are spread over a wider range as compared to ALCF no-AKI and no-progression to AKI plasma MT concentrations. A limitation of this study is the small sample size due to which this expansion in C.I. is observed and an increase in sample size is likely to reduce the C.I. and improve confidence in the ability of the cut-off to discriminate between progression and no-progression to AKI. In Kaplan-Meier analysis for time-to-AKI with the cut-off of 5.83 ng/mL plasma MT concentration, patients with MT values higher than the cut-off had a higher probability of progression to AKI by day 7 (p-value <0.0001). This suggests that plasma MT concentrations measured on day-of-admission might be a useful indicator of a progression to AKI in ACLF patients (Table 7).

MT are small (∼6 kDa) thiol-rich pleiotropic proteins that associate with multiple other proteins and participate in various processes that affect cellular stress and cell death^27-29^.The MT family of proteins is expressed by several tissues but is enriched in the liver tissue, as evidenced by the expression data extracted from the human protein atlas. Necrosis or necroptosis of hepatocytes has been shown to release MT into the circulation^20-21^. We found the expression of MT1A gene and extremely robust expression of the MT2A gene in post-mortem liver biopsies from ACLF patients, suggesting that the injured liver has high levels of inflammation characteristic of ACLF, is a source of the elevated plasma MT in ACLF. The relevance of elevated plasma MT in ACLF AKI patients, interpreted in the context of reported functions in immunomodulation and renal tissue physiology, are discussed below-

### (i) Regulation of MT expression

MT gene expression can be regulated by several factors such as toxic heavy metals, reactive oxygen species, different types of stress and cell type specific transcription factors^22^.Since heavy metal toxicity has been previously reported to be a cause of renal injury^23-24^, we assessed the levels of heavy metal ions in the plasma of ACLF patients. Detailed plasma elemental analysis for 7 different heavy metals (Cu, Zn, Ni, Cd, Hg, As, Se) ruled out the role of heavy metal poisoning as a cause for MT induction and consequent nephrotoxicity in our cohort of ACLF patients. Metal ion levels in the plasma of ACLF-AKI vs ACLF-no AKI were not found to be significantly different. However, plasma Ni levels in ACLF overall were more frequent as compared to CLD(12 of 93 ACLF patients tested has elevated plasma Ni levels) suggesting that at least in a small proportion of patients, elevated Ni levels may be a possible cause for nephrotoxicity.

However, we must also highlight that plasma trace element levels do not provide an accurate measure of tissue or cellular stores of trace elements and in these patients, the Ni levels within the lymphocytes or the kidney tissue would be an appropriate measure for these patients^20^. In most patients, however, MT induction could be a result of localized ROS generated within the liver hepatocytes due to inflammatory responses or cellular stress.

### (ii) MT and effect on reactive oxygen species

MT modulate the activity of cytotoxic ROS in two different ways- (i) intracellular MT acts as a ROS scavenger, thereby preventing cellular damage due to free radicals whereas, (ii) extracellular MT, such as the elevated plasma MT observed in our study, stimulates ROS production by innate immune cells like macrophages^29-30^. This suggests that elevated plasma MT could be promoting the innate immune pathogenesis driving ACLF, and this process may be more efficient or higher in magnitude in the fraction of ACLF patients who develop AKI. This is corroborated by several independent studies that show that ACLF patients have elevated levels of oxidative stress and ROS markers in the plasma andmacrophages and neutrophils derived from ACLF patients have higher ROS generation^30-33^.Independent studies also show that ROS accumulation is a risk factor for mortality in AKI in general^33^.This suggests that extracellular MT-induced ROS generation by macrophages and neutrophils in ACLF might be a potential mechanism by which cellular damage occurs in ACLF patients, thereby increasing the risk of AKI.

### (iii) MT-Megalin interaction in RPTEC

MT can also act as a ligand for the scavenger endocytic receptor megalin^34-35^. Human protein atlas derived expression data shows that megalin expression is maximum in renal proximal convoluted tubular epithelial cells (RPTEC) (Figure 4B; proteinatlas.org)^17^.Under normal circumstances, megalin binds to several different protein ligands in addition to MT, such as albumin, folate binding protein and vitamin D binding protein, to facilitate their re-uptake by the RPCT^19, 23^. All megalinligands compete with each other for binding ^23^. Excessive ligand binding that leads to megalin overload in RPTEC can lead to cytotoxicity and cell death ^24, 25^. In ACLF, serum albumin levels decrease due to liver failure, as reported in literature^26^. Serum albumin levels were lower in ACLF patients as compared to CLD (p-value <0.0001) in our study,as expected. We calculated MT/ Albumin ratio in order to investigate the relationship between the two liver-derived megalin ligands. In our study the MT/ Albumin (MT/ Alb) ratio was significantly elevated in ACLF patients as compared to CLD patients (p-value <0.01). ACLF AKI patients had significantly higher MT/ Alb ratiocompared to ACLF no-AKI patients (p-value<0.0001) as well as in ACLF patients who progressed to AKI vs those who did not (p-value<0.0001). These data suggest that in ACLF, the decrease in albumin concentrations is accompanied by several folds increasing MT concentrations. Therefore, MT is a major ligand for megalin binding on RPTEC in ACLF. MT has been shown to be a chemotactic factor, causing entry of leukocytes at sites of tissue stress or injury^27-28^. Therefore,the elevation of plasma MT andsubsequentincrease in its concentration at RPTECs due to megalin binding, may lead to localized tissue inflammation leading to upregulation of inflammatory markers such as TLR4 and IL-6 as reported in literature.

Putting all these observations together, we arrive at a possible pathogenic mechanism for AKI in ACLF that involves MT-interaction with immune cells as well as RPTEC, leading to excessive ROS generation and immune cell infiltration at tissue sites in ACLF patients who develop AKI. While further studies are needed to elucidate the specific molecular interactions that govern AKI development in ACLF, our study provides a basis to explore MT as a predictive biomarker of AKI in ACLF and also as a potential therapeutic target in ACLF.

## Data Availability

All data produced in the present work are contained in the manuscript

## Notes

The authors have declared that no conflict of interest exists.

**Funding:** The work carried out in this study was supported by the BioCARE grant from the Department of Biotechnology, Government of India [Grant no.BT/PR18574/BIC/101/151]. RS was supported by a fellowship from the Indian Council of Medical Research [Fellowship no.JRF-2016/HRD/LS/98/4080].

### Competing Interest Statement

The authors have declared no competing interest.

### Funding Statement

The work carried out in this study was supported by the BioCARE grant from the Department of Biotechnology, Government of India [Grant no.BT/PR18574/BIC/101/151].

### Author Declarations

This study has been approved by the Institute Ethics Committee of the All India Institute of Medical Sciences New Delhi

