## Supplementary figures and images for "Quantitative Plasma Proteomics Identifies Metallothioneins as a Marker of Acute-on-Chronic Liver Failure Associated Acute Kidney Injury"

### Supplementary figure 1

#### Slide 1
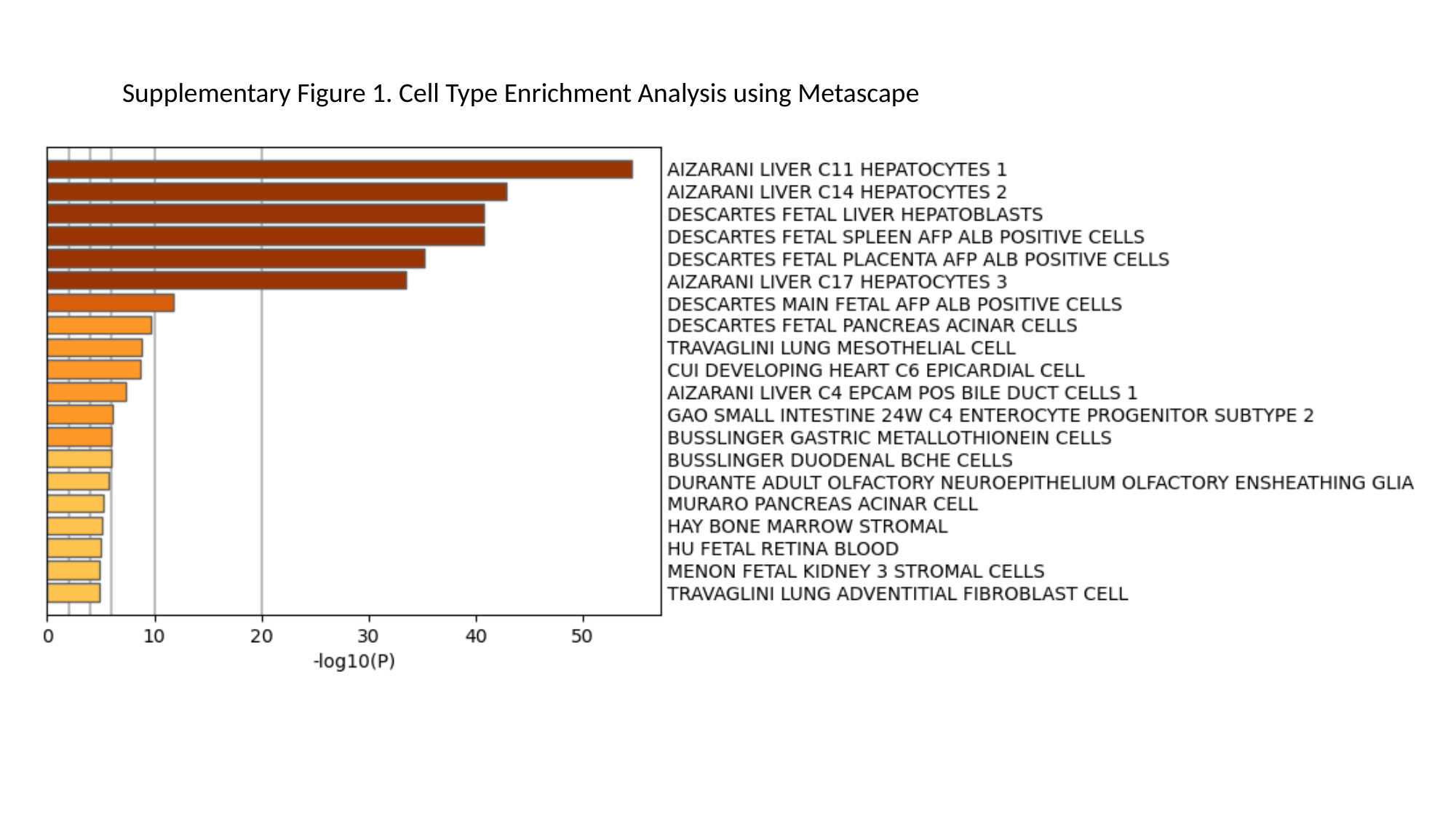

Supplementary Figure 1. Cell Type Enrichment Analysis using Metascape
