## Supplementary figure 2 for "Quantitative Plasma Proteomics Identifies Metallothioneins as a Marker of Acute-on-Chronic Liver Failure Associated Acute Kidney Injury"

### Slide 1
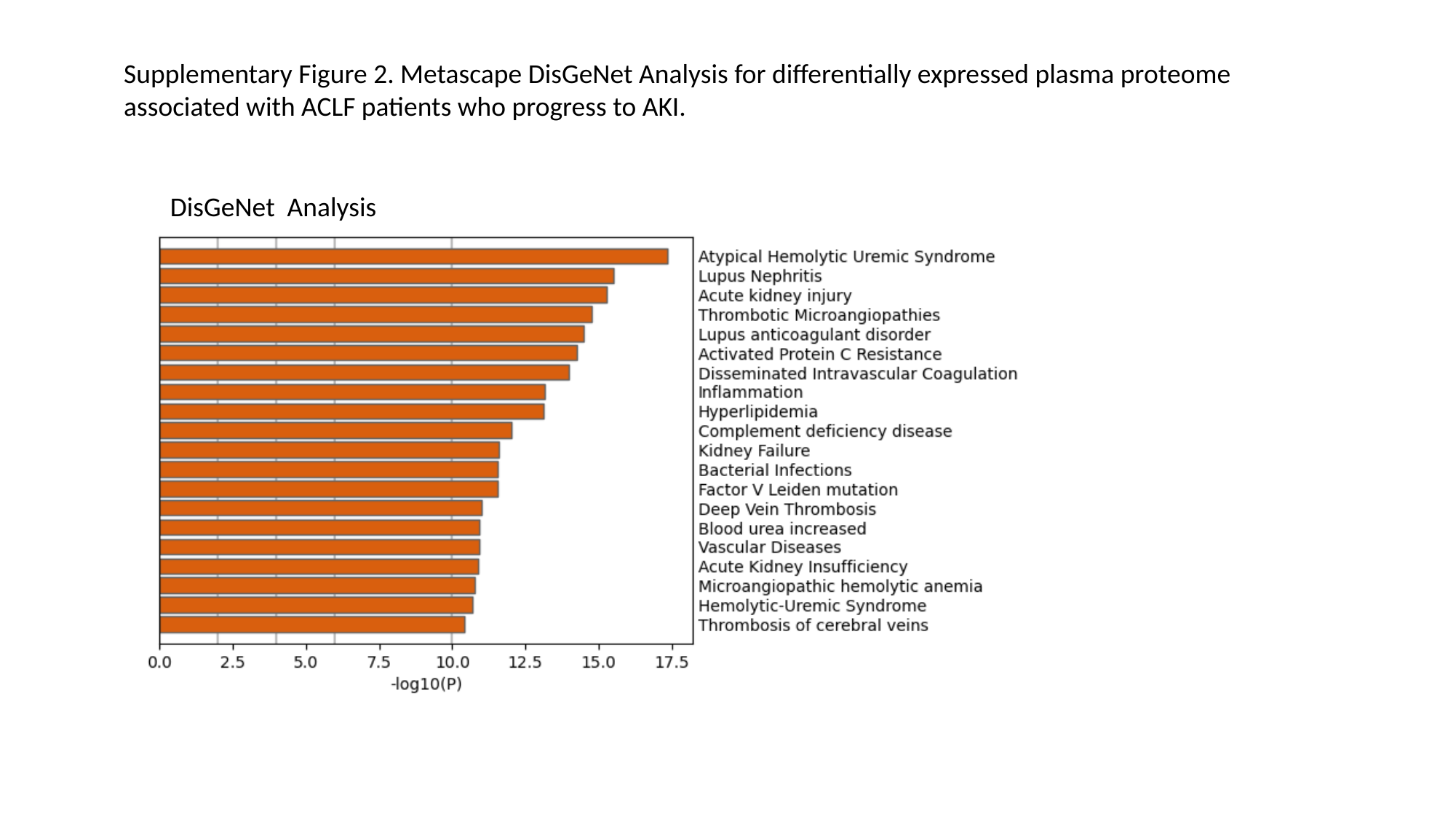

Supplementary Figure 2. Metascape DisGeNet Analysis for differentially expressed plasma proteome associated with ACLF patients who progress to AKI.
DisGeNet Analysis
