## Supplementary figure 3 for "Quantitative Plasma Proteomics Identifies Metallothioneins as a Marker of Acute-on-Chronic Liver Failure Associated Acute Kidney Injury"

### Slide 1
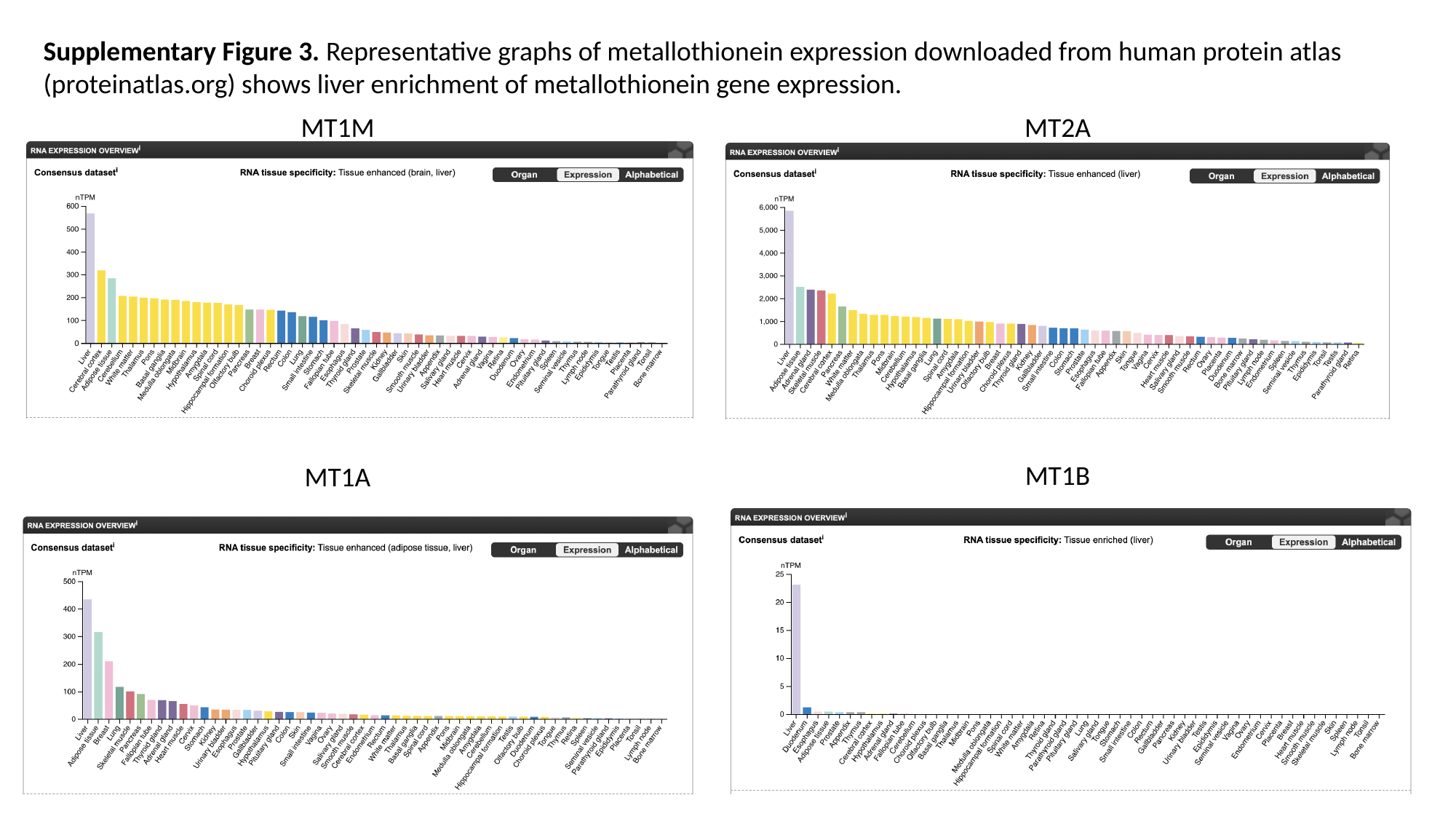

Supplementary Figure 3. Representative graphs of metallothionein expression downloaded from human protein atlas (proteinatlas.org) shows liver enrichment of metallothionein gene expression.
MT1M
MT2A
MT1B
MT1A
